# Conformal Triage for Medical Imaging AI Deployment

**DOI:** 10.1101/2024.02.09.24302543

**Authors:** Anastasios N. Angelopoulos, Stuart Pomerantz, Synho Do, Stephen Bates, Christopher P. Bridge, Daniel C. Elton, Michael H. Lev, R. Gilberto González, Michael I. Jordan, Jitendra Malik

## Abstract

**Background:** The deployment of black-box AI models in medical imaging presents significant challenges, especially in maintaining reliability across different clinical settings. These challenges are compounded by distribution shifts that can lead to failures in reproducing the accuracy attained during the AI model’s original validations.

**Method:** We introduce the conformal triage algorithm, designed to categorize patients into low-risk, high-risk, and uncertain groups within a clinical deployment setting. This method leverages a combination of a black-box AI model and conformal prediction techniques to offer statistical guarantees of predictive power for each group. The high-risk group is guaranteed to have a high positive predictive value, while the low-risk group is assured a high negative predictive value. Prediction sets are never constructed; instead, conformal techniques directly assure high accuracy in both groups, even in clinical environments different from those in which the AI model was originally trained, thereby ameliorating the challenges posed by distribution shifts. Importantly, a representative data set of exams from the testing environment is required to ensure statistical validity.

**Results:** The algorithm was tested using a head CT model previously developed by Do and colleagues [9] and a data set from Massachusetts General Hospital. The results demonstrate that the conformal triage algorithm provides reliable predictive value guarantees to a clinically significant extent, reducing the number of false positives from 233 (45%) to 8 (5%) while only abstaining from prediction on 14% of data points, even in a setting different from the training environment of the original AI model.

**Conclusions:** The conformal triage algorithm offers a promising solution to the challenge of deploying black-box AI models in medical imaging across varying clinical settings. By providing statistical guarantees of predictive value for categorized patient groups, this approach significantly enhances the reliability and utility of AI in optimizing medical imaging workflows, particularly in neuroradiology.

## 1 Introduction

AI models for medical imaging often fail to match their initial training and testing accuracy when implemented clinically [5, 6]. They struggle to *generalize*: they cannot perform as well on new data as on the data used for development. This may be due to differences in demographics—such as age, sex, acuity of presentation, and disease prevalence—as well as technical differences in hardware and protocols from the carefully curated datasets normally used to train the AI [10, 13]. The performance gap may be immediately evident, or drifting conditions may cause it to appear over time [12]. Such degradation can result in real patient harm and undermine trust in continued use of the algorithm.

The present work proposes applying an uncertainty quantification (UQ) technique called *Learn-then-Test* [2] assure *high predictive value* of a black-box AI in different clinical environments. This is achieved using recent representative imaging data from the local site. Specifically, we split patients into a low-risk group with a high negative predictive value, a high-risk group with a high positive predictive value, and an uncertain group, building on a previous approach developed at the Massachusetts General Hospital wherein neuroradiologists decide these groupings manually [3, 18]. In our new approach, these groupings are decided by a black-box machine learning algorithm, using imaging as input, in a way that endows the groupings with statistical guarantees. We call this pipeline *conformal triage*, as the triage is determined by an AI algorithm wrapped by a conformal prediction-type procedure [17, 1, 2]. Compared to existing techniques [7, 3, 18], conformal triage is post hoc (it does not require any retraining/fine-tuning), and does not make any assumptions about the form of the model or data distribution. Thus, we can provide formal guarantees of high predictive value even if there has been drift from the original training conditions.^1^

We demonstrate conformal predictive triage on a modification of the head CT model previously developed at Massachusetts General Hospital by Do and colleagues [9, 4]. The model was developed to detect intracranial hemorrhage, but when applied to real-world imaging data, it was found to also identify other major intracranial abnormalities (ICs) such as brain tumors—an unexpected, clinically useful capability. The head CT model generates a per-exam probability of a significant intracranial abnormality revealed by CT. Conformal predictive triage is then performed—critically, using a calibration set of representative site exams—to provide a statistical guarantee on both the positive predictive value (PPV) and negative predictive value (NPV) of the algorithm. Patients classified as high risk will be positive at the rate of the chosen PPV target, and similarly, those classified as low risk are guaranteed to be negative with high probability. In exchange for these guarantees, the algorithm is allowed to abstain on the most uncertain cases; the number of abstentions is determined by how stringent the PPV/NPV thresholds are.

This procedure allows the uncertainty of the model to transfer to a new local patient population, while retaining formal statistical rigor. This system is useful in neuroradiologist workflow optimization, e.g., ensuring the cases with the highest likelihood of positive findings are prioritized for diagnostic review. If properly implemented, this could benefit broader hospital operations and improve wait and discharge times.

### Contributions

Our work is the first to propose and perform a deployment and analysis of a *distribution-free* selective classification technique in the field of medicine. Methodologically, we show how to use selective classification to perform statistically valid triage. The selective classification approach, critical for downstream guarantees on the triage algorithm, distinguishes our approach from other forms of conformal prediction that have been applied in clinical medicine, e.g. [8, 14, 11], which generally output prediction sets and thus cannot be used to give triage guarantees (see [16] and [7] for helpful reviews). Our experiments provide a large-scale validation of the approach on real medical data from Mass General Brigham; to our knowledge, this is the first empirical investigation of this family of approaches in real medical imaging data not carefully curated a priori.

## 2 Methods

The standard method for tuning the performance of a binary classifier is to trade off PPV and NPV using the receiver operating characteristic (ROC) curve. Our methodology, by contrast, does not require trading off PPV and NPV; both are anchored to a pre-specified level using the calibration data, and the algorithm abstains from prediction if it is not sufficiently confident. If the calibration set is representative of future data, then our algorithm comes with formal guarantees that the PPV and NPV chosen will not fall below the desired operating thresholds. We present the mathematical details of this conformal predictive triage procedure in Section 2.1.

We applied conformal predictive triage to our IC Detection algorithm on two retrospective CT exam datasets from Massachusetts General Hospital, which we labeled ourselves; the details of these datasets are in Section 2.2. We have open-sourced the model outputs on these datasets, along with the ground truth labels, in order to facilitate the reproducibility of our results.^2^

### 2.1 Conformal predictive triage

#### Statistical setup

We use a form of selective classification to ensure the PPV and NPV are calibrated. Our statistical model is as follows. We consider a calibration data set of *n* pairs of CT scans *X*_i_ and binary labels *Y*_i_ in the set {−, +}, signifying ‘IC-negative’ and ‘IC-positive’ cases. We receive a new test scan *X*, for which there is an *unknown* binary label *Y* which we seek to predict. In this paper, we assume that the data points (*X*_1_, *Y*_1_), …, (*X*_n_, *Y*_n_), (*X, Y*) are exchangeable—invariant to ordering—which is a mathematical way to capture the intuitive notion that the calibration data is representative of the test scan.

#### Models

In order to perform the prediction, we require access to two models. The first is a deep-learning-based IC detector *g* that takes the images from a head CT exam as input and outputs a probability of IC for each slice of the scan. The second is an aggregator function *h* that maps the output of *g* to a number between 0 and 1 signifying the estimated probability of IC in the entire brain. The IC classifier *g* is the ML model developed by Do and colleagues in [9, 4], and we experiment with different functions *h*, including isotonic and logistic regressions; see Section 3. For convenience, we give the name *f* = *h* ∘ *g* to the chain of models (i.e., *f* (*x*) = *h*(*g*(*x*))).

#### Formal Goal

The goal is to achieve both a high PPV and a high NPV by allowing the model to abstain on the cases that exceed the uncertainty threshold, i.e., for which the statistical guarantee for PPV or NPV can not be met. Towards that end, we introduce a parameter λ that controls how many abstentions (non-predictions) are made. In particular, we set *Ŷ* (*x*), the prediction of *Y*, to be

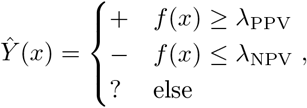

and we let λ = (*λ*_FPR_, *λ*_FNR_) denote the pair of parameters. The goal is to select a data-driven value 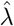 satisfying, with high probability, that the false positive rate (FPR) and the false negative rate (FNR) are controlled:

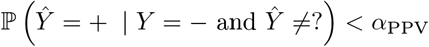

and

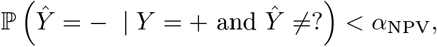

for some user-chosen parameters *α*_PPV_ and *α*_NPV_. The parameter 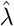 will be selected based on a calibration dataset that is assumed to be representative of future data points. We want to guarantee, without any further assumptions on the form of the data distribution or the model *f*, that the PPV and NPV are greater than or equal to 1 − *α*_PPV_ and 1 − *α*_NPV_, respectively. This assumption-light statistical approach allows our method to apply to any model and data distribution, as long as an appropriate calibration set is available.

#### Method

We build upon the Learn then Test variant of conformal prediction developed in [2]. The method constructs discrete grid, Λ = {0, 1*/m*, …, (*m* − 1)*/m*, 1} from 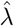 is selected. For false positive rate control,

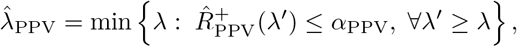

where

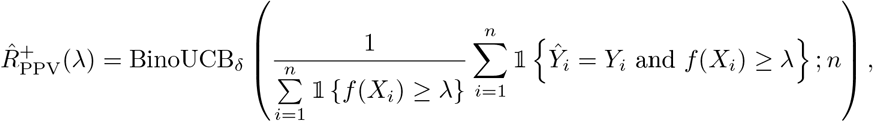

and the function BinoUCB_δ_ is the upper end of the binomial confidence interval constructed at level 1 − *δ*. The second coordinate, 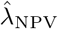, is constructed analogously. The user can elect to apply a Bonferroni correction (i.e., to choose both coordinates with level *δ/*2) to achieve a simultaneous guarantee for PPV and NPV. This strategy is generally statistically tight.

### 2.2 Description of Data Sets

We perform experiments with two data sets of head CT scans collected at Massachusetts General Hospital along with a pre-trained, pre-existing intracranial hemorrhage detector *f* built on historical data from the hospital system; details are referred to the prior work [9, 4]. In both datasets, the feature *X*_i_ is a CT scan with a variable number of slices. The label *Y*_i_ is binary, corresponding to an indicator {−, +} of an IC. The first data set 𝒟_1_ is a painstakingly curated data set consisting of 827 consecutive CT exams which were scrutinized by a senior, board-certified neuroradiologist with over 30 years of experience to produce the labels *Y*_i_—in this sense, the dataset truly contains gold-standard labels. The second data set 𝒟_2_ contains 9122 consecutive CT exams whose ground truth labels were extracted by a regular-expression-based pattern-matching algorithm on the radiology report text, which was then manually verified by a senior neuroradioloigst (SRP) with greater than 20 years experience. Each data point also contains a binary indicator of diagnostic indeterminacy—cases where the neuroradiologist that originally read the scan was not certain of the disease state—and also an indicator of parsing indeterminacy, i.e., cases where the original neuroradiologist may have understood the disease state, but did not communicate it in a clear way in their notes. The data set 𝒟_2_ is of reasonably high quality, representing the standard of care that patients receive at a hospital; not all scans are read carefully by experienced neuroradiologists, and thus there may be more errors in these labels than in 𝒟_1_.

## 3. Experiments

### Setup

We use several methods of regression. To estimate the function *h* which is chained with the original slice-wise risk predictor *g*, we evaluate the use of isotonic regression, a logistic regression model, and a simple thresholding classifier. All three algorithms work reasonably well, and we focus on the thresholding classifier for the sake of simplicity. In the figures, models are trained using 3000 data points from 𝒟_2_ and evaluated on the remaining data points from 𝒟_2_ as well as those from 𝒟_1_. Below are given representative quantitative results on both 𝒟_1_ and the validation split of 𝒟_2_, and also plots of the accuracy as a function of *λ* in order to understand the qualitative behavior of the procedure and its tightness (in general, it is slightly conservative). It should be noted that the choice of 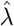 is optimized on 𝒟_2_, so the data in 𝒟_1_ is subject to a shifting distribution. On 𝒟_2_, we also report results on the subset of data points without any diagnostic or parsing indeterminacies (this subset is labeled ‘no ID’).

Our main point of comparison is a heuristically derived set of thresholds manually designed by the expert neuroradiologist. The neuroradiologist developed a hand-designed thresholding rule on a consecutive series of ED head CTs. It was tested on a subsequent 1000 consecutive cases from the ED. The rule is that if *h*(*x*) assigns any three slices to have an estimated probability of at least 0.9, then we set *f* (*x*) = +. If not, and if additionally there are no more than a single slice above 0.6, then *f* (*x*) = −. Otherwise, *f* (*x*) is set to equal ?. This is a strong baseline in-distribution on 𝒟_1_ (the in-distribution data), but it is limited in reproducibility and generalizability to different and changing patient populations. Our machine learning approach can achieve a similar PPV/NPV tradeoff, while remaining tunable to different PPV/NPV set points. It is a scalable and repeatable procedure for calibrating AI algorithms in real hospital scenarios that avoids the need for a neuroradiologist to hand-design such algorithms.

The plots include enough information to reconstruct the output of our procedure for any choices of *α*_PPV_ and *α*_NPV_. As a visual aid, we plot as gray lines the value of 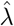 chosen with *α*_PPV_ = 0.3 and *α*_NPV_ = 0.05. This mirrors the use case of triage discussed in the introduction: the NPV threshold is substantially more stringent so as to avoid missing positive cases. Intuitively, the ramification of this choice is that the procedure will be more aggressive in identifying positive cases than negative ones.

### Top-line result

Conformal triage effectively calibrates the previously developed model to a *current* set of patient exams, enabling high-confidence PPV and NPV compared to the original model, without the need for retraining and an acceptable rate of abstention (non-prediction). The exact numbers depend on the desired tradeoff between abstention rate and accuracy. On the most extreme end, the calibrated isotonic regression procedure can provide a formal PPV guarantee of at least 95% and NPV of at least 95%, with only 4.9% of data points abstained; see the third row from the bottom of Table 2. Results on this extreme end tend to be unstable due to the small number of samples, so in our plots, we calibrate to a PPV of 90% and an NPV of 95% using the logistic regression procedure, leading to an abstention rate of 14% on the validation data. By standard ROC analysis, an NPV of 95% would yield PPV of 55%, a decrease of 40 percentage points. On our data, this is a decrease from to 233 to 8 false positives—a clinically significant result.

#### Comparison of different regression methods

Tables 1 and 2 include results on all the regression algorithms. The baseline performance of the model using the heuristically derived thresholds is included in the caption of the tables. Our algorithm matches or exceeds the performance of the hand-designed algorithm. It is important to notice that on 𝒟_1_ the hand-designed algorithm has a PPV of 0.92, while on 𝒟_2_ the PPV is 0.7. This is a large drop in performance that is caused by the distribution shift in the population of the two datasets, and it is exactly the sort of situation we want to solve with our methodology. The calibrated version achieves the desired statistical guarantee of PPV and NPV for all settings of *α*_PPV_ and *α*_NPV_.

**Table 1:**
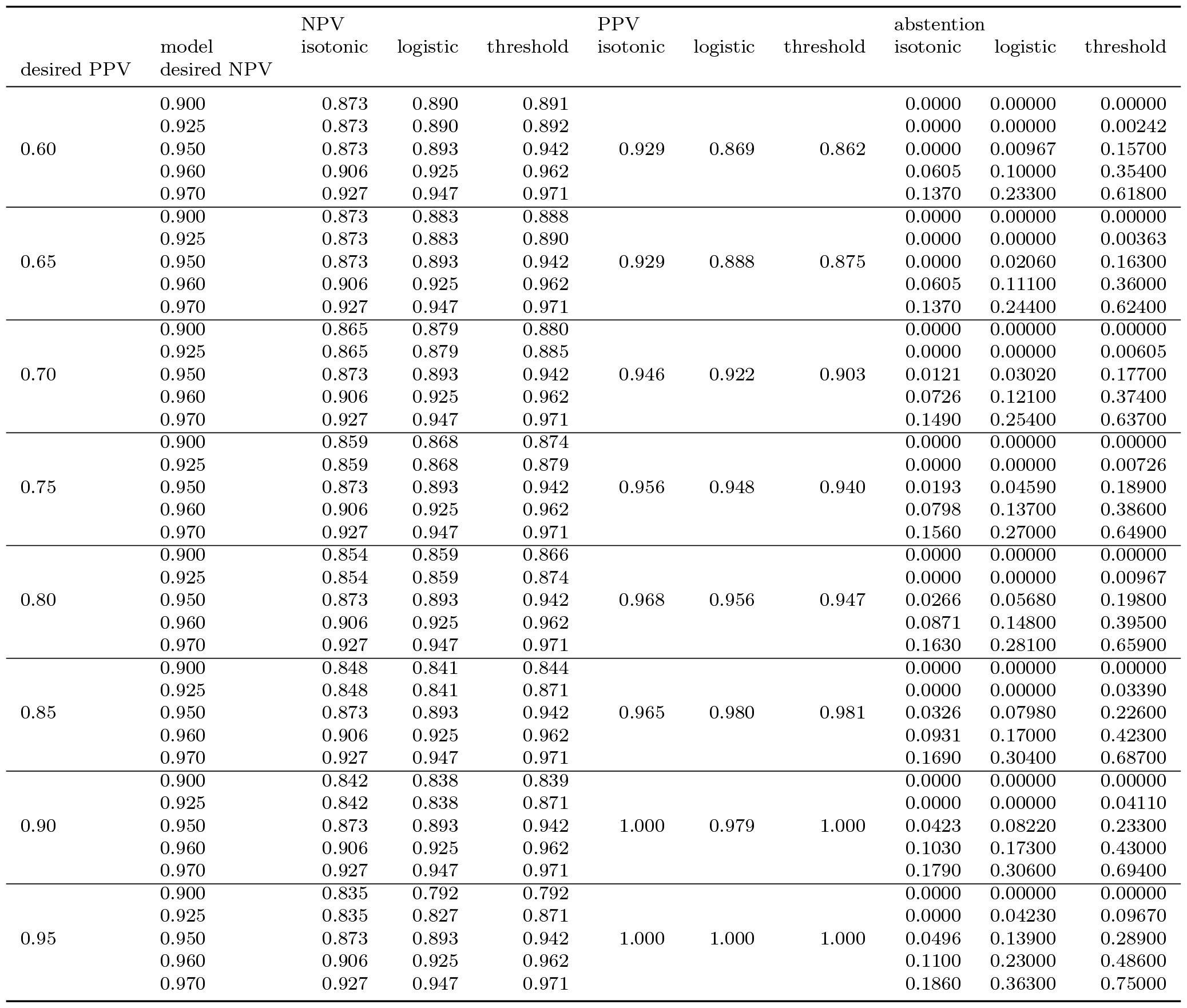
Numerical results of PPV, NPV, and abstention (non-prediction) rate as a function of model type and desired PPV and NPV guarantee level. Results are computed on 𝒟_1_. The baseline hand-designed rule achieves a PPV of 0.92, an NPV of 0.94, and an abstention rate of 0.17.

| desired PPV | model<br>desired NPV | NPV |  |  | PPV |  |  | abstention |  |  |
| --- | --- | --- | --- | --- | --- | --- | --- | --- | --- | --- |
|  |  | isotonic | logistic | threshold | isotonic | logistic | threshold | isotonic | logistic | threshold |
| 0.60 | 0.900 | 0.873 | 0.890 | 0.891 | 0.929 | 0.869 | 0.862 | 0.0000 | 0.00000 | 0.00000 |
|  | 0.925 | 0.873 | 0.890 | 0.892 |  |  |  | 0.0000 | 0.00000 | 0.00242 |
|  | 0.950 | 0.873 | 0.893 | 0.942 |  |  |  | 0.0000 | 0.00967 | 0.15700 |
|  | 0.960 | 0.906 | 0.925 | 0.962 |  |  |  | 0.0605 | 0.10000 | 0.35400 |
|  | 0.970 | 0.927 | 0.947 | 0.971 |  |  |  | 0.1370 | 0.23300 | 0.61800 |
| 0.65 | 0.900 | 0.873 | 0.883 | 0.888 | 0.929 | 0.888 | 0.875 | 0.0000 | 0.00000 | 0.00000 |
|  | 0.925 | 0.873 | 0.883 | 0.890 |  |  |  | 0.0000 | 0.00000 | 0.00363 |
|  | 0.950 | 0.873 | 0.893 | 0.942 |  |  |  | 0.0000 | 0.02060 | 0.16300 |
|  | 0.960 | 0.906 | 0.925 | 0.962 |  |  |  | 0.0605 | 0.11100 | 0.36000 |
|  | 0.970 | 0.927 | 0.947 | 0.971 |  |  |  | 0.1370 | 0.24400 | 0.62400 |
| 0.70 | 0.900 | 0.865 | 0.879 | 0.880 | 0.946 | 0.922 | 0.903 | 0.0000 | 0.00000 | 0.00000 |
|  | 0.925 | 0.865 | 0.879 | 0.885 |  |  |  | 0.0000 | 0.00000 | 0.00605 |
|  | 0.950 | 0.873 | 0.893 | 0.942 |  |  |  | 0.0121 | 0.03020 | 0.17700 |
|  | 0.960 | 0.906 | 0.925 | 0.962 |  |  |  | 0.0726 | 0.12100 | 0.37400 |
|  | 0.970 | 0.927 | 0.947 | 0.971 |  |  |  | 0.1490 | 0.25400 | 0.63700 |
| 0.75 | 0.900 | 0.859 | 0.868 | 0.874 | 0.956 | 0.948 | 0.940 | 0.0000 | 0.00000 | 0.00000 |
|  | 0.925 | 0.859 | 0.868 | 0.879 |  |  |  | 0.0000 | 0.00000 | 0.00726 |
|  | 0.950 | 0.873 | 0.893 | 0.942 |  |  |  | 0.0193 | 0.04590 | 0.18900 |
|  | 0.960 | 0.906 | 0.925 | 0.962 |  |  |  | 0.0798 | 0.13700 | 0.38600 |
|  | 0.970 | 0.927 | 0.947 | 0.971 |  |  |  | 0.1560 | 0.27000 | 0.64900 |
| 0.80 | 0.900 | 0.854 | 0.859 | 0.866 | 0.968 | 0.956 | 0.947 | 0.0000 | 0.00000 | 0.00000 |
|  | 0.925 | 0.854 | 0.859 | 0.874 |  |  |  | 0.0000 | 0.00000 | 0.00967 |
|  | 0.950 | 0.873 | 0.893 | 0.942 |  |  |  | 0.0266 | 0.05680 | 0.19800 |
|  | 0.960 | 0.906 | 0.925 | 0.962 |  |  |  | 0.0871 | 0.14800 | 0.39500 |
|  | 0.970 | 0.927 | 0.947 | 0.971 |  |  |  | 0.1630 | 0.28100 | 0.65900 |
| 0.85 | 0.900 | 0.848 | 0.841 | 0.844 | 0.965 | 0.980 | 0.981 | 0.0000 | 0.00000 | 0.00000 |
|  | 0.925 | 0.848 | 0.841 | 0.871 |  |  |  | 0.0000 | 0.00000 | 0.03390 |
|  | 0.950 | 0.873 | 0.893 | 0.942 |  |  |  | 0.0326 | 0.07980 | 0.22600 |
|  | 0.960 | 0.906 | 0.925 | 0.962 |  |  |  | 0.0931 | 0.17000 | 0.42300 |
|  | 0.970 | 0.927 | 0.947 | 0.971 |  |  |  | 0.1690 | 0.30400 | 0.68700 |
| 0.90 | 0.900 | 0.842 | 0.838 | 0.839 | 1.000 | 0.979 | 1.000 | 0.0000 | 0.00000 | 0.00000 |
|  | 0.925 | 0.842 | 0.838 | 0.871 |  |  |  | 0.0000 | 0.00000 | 0.04110 |
|  | 0.950 | 0.873 | 0.893 | 0.942 |  |  |  | 0.0423 | 0.08220 | 0.23300 |
|  | 0.960 | 0.906 | 0.925 | 0.962 |  |  |  | 0.1030 | 0.17300 | 0.43000 |
|  | 0.970 | 0.927 | 0.947 | 0.971 |  |  |  | 0.1790 | 0.30600 | 0.69400 |
| 0.95 | 0.900 | 0.835 | 0.792 | 0.792 | 1.000 | 1.000 | 1.000 | 0.0000 | 0.00000 | 0.00000 |
|  | 0.925 | 0.835 | 0.827 | 0.871 |  |  |  | 0.0000 | 0.04230 | 0.09670 |
|  | 0.950 | 0.873 | 0.893 | 0.942 |  |  |  | 0.0496 | 0.13900 | 0.28900 |
|  | 0.960 | 0.906 | 0.925 | 0.962 |  |  |  | 0.1100 | 0.23000 | 0.48600 |
|  | 0.970 | 0.927 | 0.947 | 0.971 |  |  |  | 0.1860 | 0.36300 | 0.75000 |

**Table 2:** Numerical results of PPV, NPV, and abstention (non-prediction) rate as a function of model type and desired PPV and NPV guarantee level. Results are computed on 𝒟_2_. The baseline hand-designed rule achieves a PPV of 0.7, an NPV of 0.96, and an abstention rate of 0.14.

| desired PPV | model<br>desired NPV | NPV |  |  | PPV |  |  | abstention |  |  |
| --- | --- | --- | --- | --- | --- | --- | --- | --- | --- | --- |
|  |  | isotonic | logistic | threshold | isotonic | logistic | threshold | isotonic | logistic | threshold |
| 0.60 | 0.900 | 0.942 | 0.947 | 0.948 | 0.703 | 0.625 | 0.646 | 0.000 | 0.000 | 0.000 |
|  | 0.925 | 0.942 | 0.947 | 0.949 |  |  |  | 0.000 | 0.000 | 0.003 |
|  | 0.950 | 0.942 | 0.947 | 0.963 |  |  |  | 0.000 | 0.003 | 0.146 |
|  | 0.960 | 0.952 | 0.956 | 0.970 |  |  |  | 0.044 | 0.071 | 0.316 |
|  | 0.970 | 0.964 | 0.967 | 0.987 |  |  |  | 0.107 | 0.193 | 0.610 |
| 0.65 | 0.900 | 0.942 | 0.943 | 0.943 | 0.703 | 0.687 | 0.657 | 0.000 | 0.000 | 0.000 |
|  | 0.925 | 0.942 | 0.943 | 0.945 |  |  |  | 0.000 | 0.000 | 0.006 |
|  | 0.950 | 0.942 | 0.947 | 0.963 |  |  |  | 0.000 | 0.016 | 0.155 |
|  | 0.960 | 0.952 | 0.956 | 0.970 |  |  |  | 0.044 | 0.084 | 0.325 |
|  | 0.970 | 0.964 | 0.967 | 0.987 |  |  |  | 0.107 | 0.206 | 0.619 |
| 0.70 | 0.900 | 0.938 | 0.941 | 0.940 | 0.714 | 0.741 | 0.717 | 0.000 | 0.000 | 0.000 |
|  | 0.925 | 0.938 | 0.941 | 0.942 |  |  |  | 0.000 | 0.000 | 0.007 |
|  | 0.950 | 0.942 | 0.947 | 0.963 |  |  |  | 0.008 | 0.025 | 0.165 |
|  | 0.960 | 0.952 | 0.956 | 0.970 |  |  |  | 0.052 | 0.093 | 0.335 |
|  | 0.970 | 0.964 | 0.967 | 0.987 |  |  |  | 0.115 | 0.215 | 0.629 |
| 0.75 | 0.900 | 0.937 | 0.940 | 0.941 | 0.780 | 0.750 | 0.754 | 0.000 | 0.000 | 0.000 |
|  | 0.925 | 0.937 | 0.940 | 0.942 |  |  |  | 0.000 | 0.000 | 0.008 |
|  | 0.950 | 0.942 | 0.947 | 0.963 |  |  |  | 0.014 | 0.027 | 0.168 |
|  | 0.960 | 0.952 | 0.956 | 0.970 |  |  |  | 0.058 | 0.095 | 0.338 |
|  | 0.970 | 0.964 | 0.967 | 0.987 |  |  |  | 0.121 | 0.217 | 0.632 |
| 0.80 | 0.900 | 0.934 | 0.938 | 0.939 | 0.800 | 0.769 | 0.788 | 0.000 | 0.000 | 0.000 |
|  | 0.925 | 0.934 | 0.938 | 0.940 |  |  |  | 0.000 | 0.000 | 0.010 |
|  | 0.950 | 0.942 | 0.947 | 0.963 |  |  |  | 0.019 | 0.031 | 0.173 |
|  | 0.960 | 0.952 | 0.956 | 0.970 |  |  |  | 0.063 | 0.099 | 0.343 |
|  | 0.970 | 0.964 | 0.967 | 0.987 |  |  |  | 0.126 | 0.221 | 0.637 |
| 0.85 | 0.900 | 0.933 | 0.934 | 0.933 | 0.875 | 0.818 | 0.833 | 0.000 | 0.000 | 0.000 |
|  | 0.925 | 0.933 | 0.934 | 0.939 |  |  |  | 0.000 | 0.000 | 0.018 |
|  | 0.950 | 0.942 | 0.947 | 0.963 |  |  |  | 0.024 | 0.039 | 0.183 |
|  | 0.960 | 0.952 | 0.956 | 0.970 |  |  |  | 0.068 | 0.107 | 0.353 |
|  | 0.970 | 0.964 | 0.967 | 0.987 |  |  |  | 0.131 | 0.229 | 0.647 |
| 0.90 | 0.900 | 0.929 | 0.931 | 0.932 | 0.909 | 0.914 | 0.917 | 0.000 | 0.000 | 0.000 |
|  | 0.925 | 0.929 | 0.931 | 0.939 |  |  |  | 0.000 | 0.000 | 0.024 |
|  | 0.950 | 0.942 | 0.947 | 0.963 |  |  |  | 0.031 | 0.048 | 0.189 |
|  | 0.960 | 0.952 | 0.956 | 0.970 |  |  |  | 0.075 | 0.116 | 0.359 |
|  | 0.970 | 0.964 | 0.967 | 0.987 |  |  |  | 0.138 | 0.238 | 0.653 |
| 0.95 | 0.900 | 0.927 | 0.901 | 0.901 | 0.933 | 1.000 | 1.000 | 0.000 | 0.000 | 0.000 |
|  | 0.925 | 0.927 | 0.925 | 0.939 |  |  |  | 0.000 | 0.027 | 0.060 |
|  | 0.950 | 0.942 | 0.947 | 0.963 |  |  |  | 0.034 | 0.083 | 0.225 |
|  | 0.960 | 0.952 | 0.956 | 0.970 |  |  |  | 0.078 | 0.151 | 0.395 |
|  | 0.970 | 0.964 | 0.967 | 0.987 |  |  |  | 0.141 | 0.273 | 0.689 |

#### Tracing the PPV and NPV as a function of *λ*

We plot the empirical PPV and NPV on test data as a function of *λ* in Figures 1 and 2. The figures indicate that the NPV and PPV are controlled at the desired level on 𝒟_2_. The control is more conservative on the PPV side than for the NPV side, as there are fewer positive data points; the conservativeness would go away with the use of more data. On the other hand, on 𝒟_1_, the PPV is controlled, but the NPV is not. The root cause is that the calibration data has more negatives than 𝒟_1_, since 𝒟_1_ is higher quality and is less likely to have positives that are missed. This is further evidence of the imperative to collect good ground-truth data and a calibration set that represents the future patient population to ensure the proper error rate. Finally, we note that although the PPV is controlled, it is quite conservative; this is because positive cases are relatively rare in our data, so there is not enough information to calibrate the PPV to a very stringent level.

**Figure 1:**
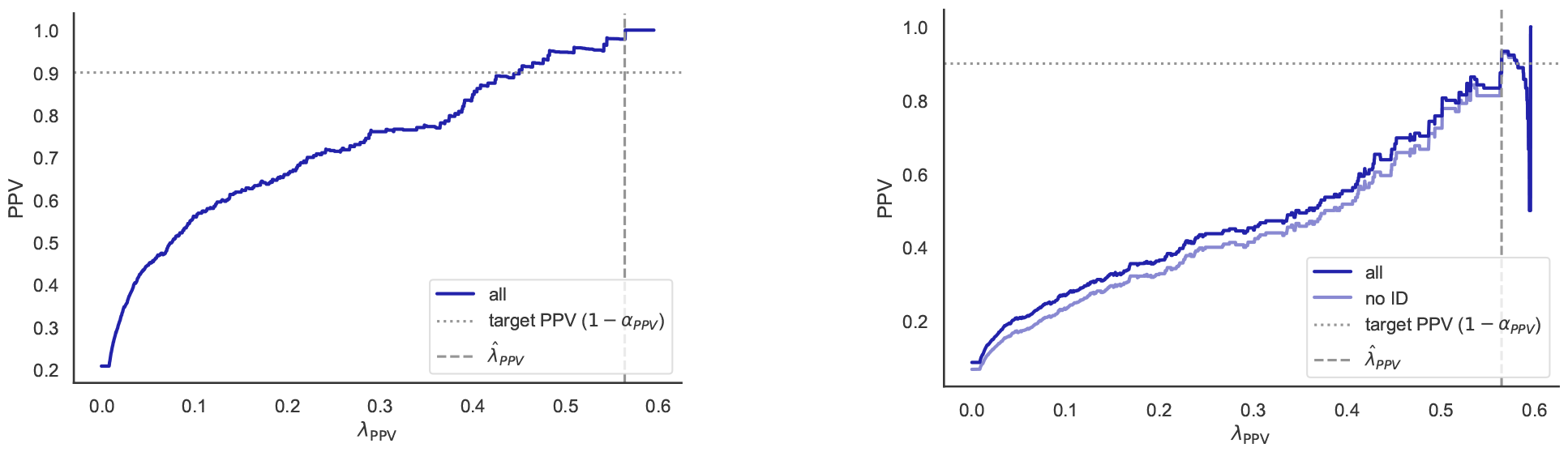
PPV as a function of the threshold. The horizontal axis is the parameter *λ*_PPV_. The vertical axis is the PPV, i.e., the fraction of the scans labeled + that are indeed +. The gray dotted line indicates the target PPV, which is guaranteed to hold with probability at least 1 − *α*_PPV_. The left plot is computed on 𝒟_1_ and the right is on 𝒟_2_.

**Figure 2:**
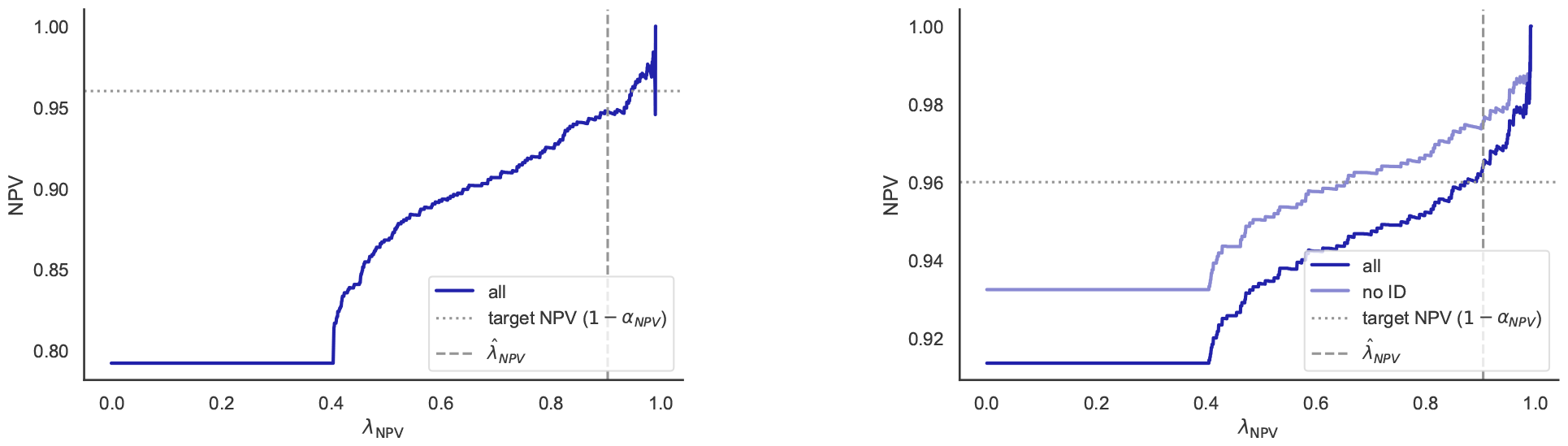
NPV as a function of the threshold. The horizontal axis is the parameter *λ*_NPV_. The vertical axis is the NPV, i.e., the fraction of the scans labeled − that are indeed −. The left plot is computed on 𝒟_1_ and the right is on 𝒟_2_.

#### Evaluating the True Positive Rate (TPR) and True Negative Rate (TNR)

Next we look at the TPR and TNR to understand how many positive and negative cases are assigned predictions (as opposed to the abstention/non-prediction category, ‘?’). The PPV often trades off with the TPR, as increasing the number of abstentions improves the former and hurts the latter (since fewer positive cases are recovered). The analogous principle holds for negative cases. Figures 3 and 4 illustrate this tradeoff.

**Figure 3:**
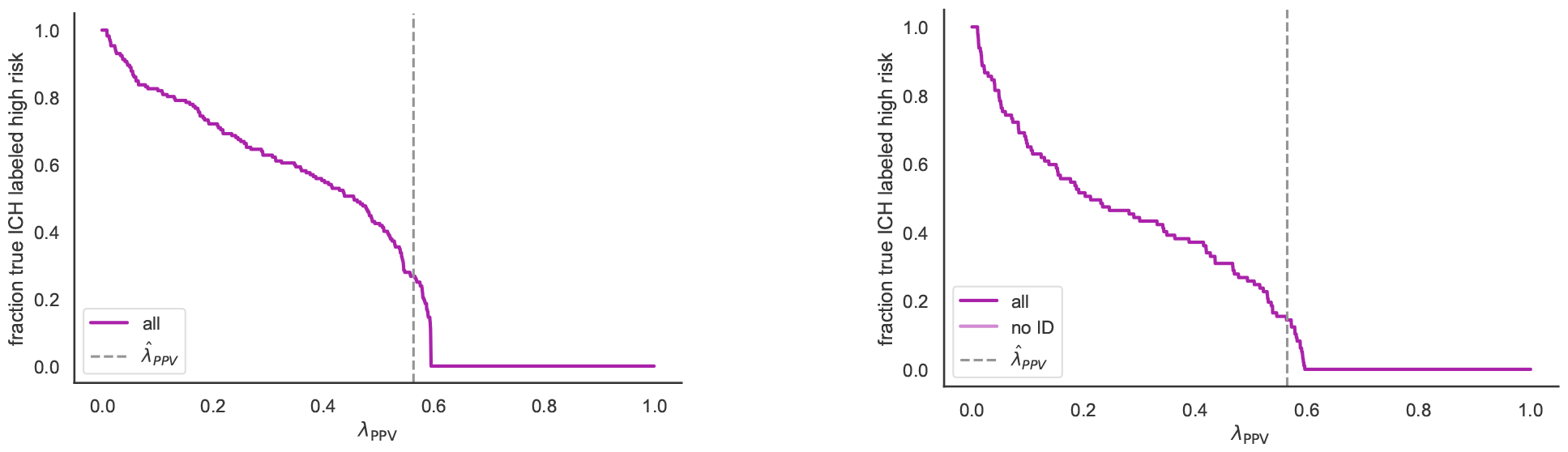
TPR as a function of the threshold. The horizontal axis is the parameter *λ*_PPV_. The vertical axis is the TPR, i.e., the fraction of actually positive scans labeled positive. The left plot is computed on 𝒟_1_ and the right is on 𝒟_2_.

**Figure 4:**
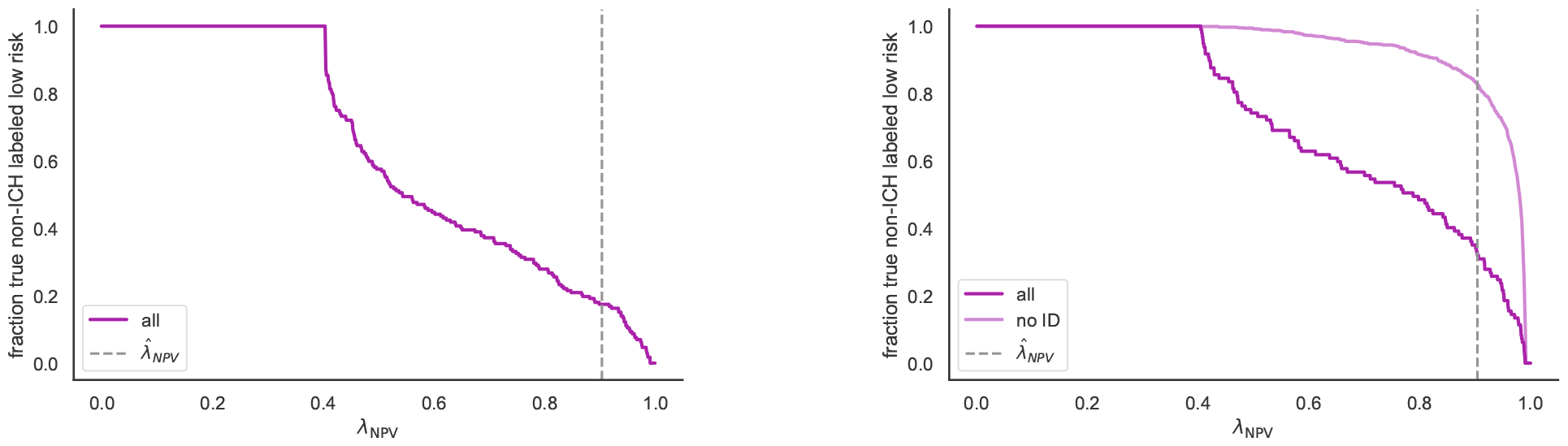
TNR as a function of the threshold. The horizontal axis is the parameter *λ*_NPV_. The vertical axis is the TNR, i.e., the fraction of actually negative scans labeled negative. The left plot is computed on 𝒟_1_ and the right is on 𝒟_2_.

#### Checking the marginal proportion of + and – cases

It is of interest to check what fraction of the total cases are assigned to the classes + and – as a function of *λ*—this is called the *marginal proportion of positive (or negative) cases*. Of course, setting *λ*_PPV_ = 0 would result in all cases being assigned to the class +, and then the marginal proportion of positives would be 1. This is clearly undesirable because it means a large false positive rate. However, it may be necessary to over-predict the class + in order to achieve the safety guarantee, due to the stringent choice of *α*_NPV_; this is what the data in Figures 5 and 6 indicate. Meanwhile, the negative class is slightly under-predicted as *α*_PPV_ is less stringent. These effects are not particularly extreme, but are tunable by changing the *α* parameters, as one can witness by examining the shape of the curves.

**Figure 5:**
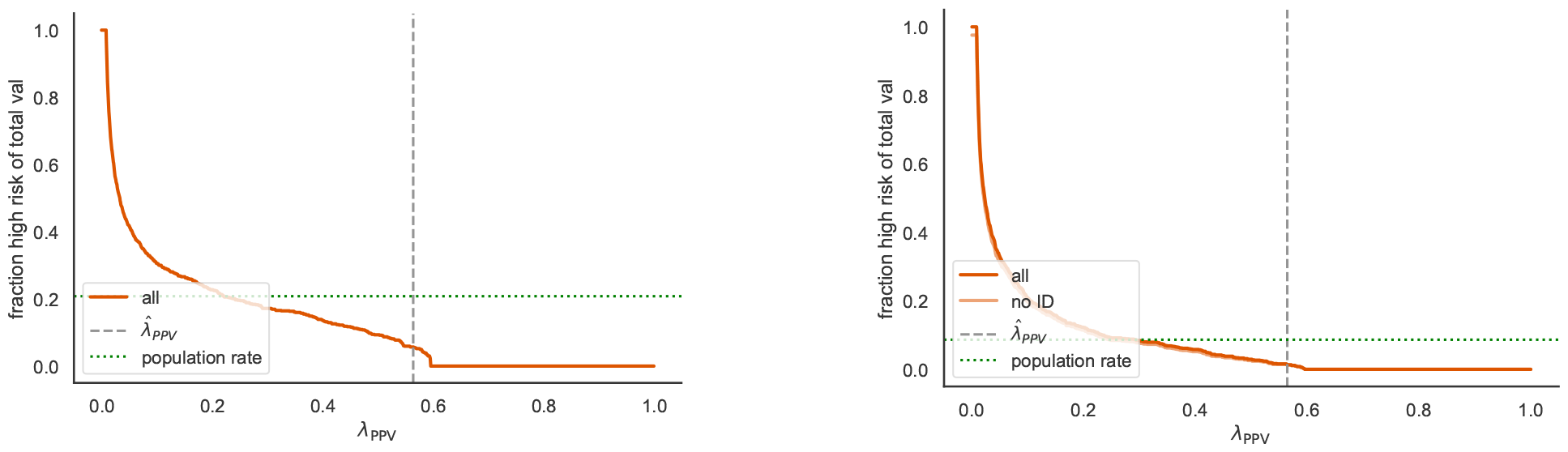
Marginal proportion of cases assigned + as a function of the threshold. The horizontal axis is the parameter *λ*_PPV_. The vertical axis is the fraction of cases assigned to the class +. The left plot is computed on 𝒟_1_ and the right is on 𝒟_2_.

**Figure 6:**
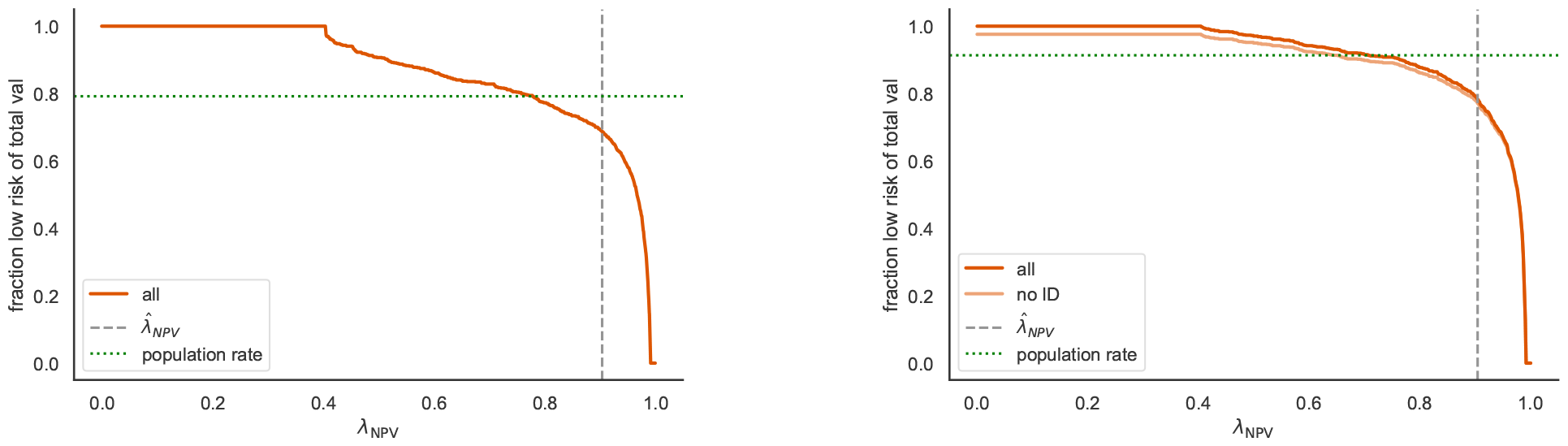
Marginal proportion of cases assigned - as a function of the threshold. The horizontal axis is the parameter *λ*_NPV_. The vertical axis is the fraction of cases assigned to the class -. In green is shown the true population fraction of - cases. The left plot is computed on 𝒟_1_ and the right is on 𝒟_2_.

## 4 Discussion

Conformal uncertainty quantification provides a non-heuristic approach to accurate medical AI in varying conditions, yielding better decision-making and risk management. This provides guarantees of reliable performance using an automated algorithm that can be re-run as the data distribution changes, even if the base model cannot be retrained. The requirement is a small set of exams representative of the population. Different strategies for tuning our PPV and NPV guarantees enable different clinical use-cases, as described below.

### Rule-out triage

This approach prioritizes minimizing the false-negative rates to effectively ‘rule out’ non-IC cases. Setting *α*_*NPV*_ to be small leads to a high accuracy in the negative group. The implication is that most of the actual negative cases are correctly identified, and these can be prepared for discharge, while safely directing hospital resources towards the more urgent cases.

### Rule-in triage

This strategy focuses on minimizing false-positive rates to ‘rule in’ potential IC cases. This involves setting *α*_*PPV*_ to be quite small, leading to a high accuracy in the positive group. The implication is that the diagnosed positive cases are actual positives, minimizing the resources spent on false alarms, and ensuring that neuroradiologists can read the most urgent scans quickly.

Both the rule-out and rule-in triage approaches can be achieved with only *one* of the PPV and NPV control parameters, i.e., one of the coordinates of *λ*. This is akin to a more rigorous form of ROC curve analysis. However, our approach also allows for simultaneous control of PPV and NPV, similar to picking two points on the ROC curve simultaneously.

### Real-time triage

This approach assures *both* high specificity and sensitivity, providing real-time insights that are neither too conservative nor too liberal. It involves picking both *α*_PPV_ and *α*_NPV_ to achieve both forms of control simultaneously, while leaving the middle group as uncertain. It facilitates more dynamic decision-making and can adapt to changing circumstances and data distributions.

The algorithms that we have presented are feasible in practice, do not require manual intervention, and are straightforward to implement. The data requirements are limited; the *n* required for calibration is not very large unless the levels *α*_PPV_ and *α*_NPV_ are quite stringent. Importantly, the approach is post hoc, meaning it can work with any AI model with no requirements on the training/development of that model. This makes it an attractive tool for deployment of medical imaging AI, and potentially also as a tool for regulators [15] to give basic guidelines that the models follow rigorous validation protocols.

## Data Availability

All data produced are available online at https://github.com/aangelopoulos/conformal-triage.

## Footnotes

1 Importantly, the guarantee assumes access to a small amount of representative calibration data from the new domain.

2 Code available at https://github.com/aangelopoulos/conformal-triage.

